# Is India heading towards a new high? : An optimistic approach to estimate ending life-cycle and cumulative cases by the end of the major COVID-19 pandemic wave in India and some of its states

**DOI:** 10.1101/2020.05.30.20117440

**Authors:** P. Gupta, K. K. Sharma, S.D. Joshi, S. Goyal

## Abstract

Projecting the COVID-19 curve parameters such as ending-lifecycle and cumulative cases are helpful in guiding the policy makers to mitigate the outbreak. However, overestimating these parameters may put the public and policy makers in a muddle. In this paper, an optimistic scenario is simulated, wherein the dynamics of the COVID-19 curve is allowed to spread to such an extent that the projections of the COVID-19 parameters do not take excessively high values. Based on this scenario, the ending life-cycle and cumulative cases for India and some of its states, are predicted. Our study, suggests that the fall of the peak amplitude (95%) of the major COVID-19 wave in India may take place by the 8^th^ of September 2020 with a total count of 655000 cases. Simulation results, also indicate that Maharashtra, Tamil Nadu, Delhi, Gujarat, Uttar Pradesh, Bihar, Madhya Pradesh, and Rajasthan may end up with 263700, 18140, 50600, 21130, 24420, 44170, 27080, and 28200 cumulative cases respectively.

## 1. Introduction

The Chinese city, Wuhan first turned into an epicentre of disease due to a severe acute respiratory syndrome coronavirus 2 (SARS-CoV-2), in December 2019 [1-3]. The situation started becoming grim around the world when travellers started roaming from China to the rest of the countries. World health organization declared the Coronavirus-19 disease (COVID-19) as a pandemic on 11^th^ March 2020 [4]. In the absence of a vaccine and specific treatment, most of the countries adopted the social distancing norm as one of the major policy interventions to mitigate the COVID-19 outbreak. However, estimating the COVID-19 dynamics and parameters such as cumulative cases, deaths, flattening of the curve, and ending life-cycle, at regular intervals are also vital for shaping these mitigation strategies in the right direction. Some of the taskforces rapidly disseminated information regarding these parameters to make public and governments aware regarding important facts about this disease [4-6]

Susceptible-Infected-Recovered (SIR) and its variants are established and the most common models to estimate the epidemiological disease dynamics [7-10]. Recently, many researchers tried these methods to estimate the disease dynamics of COVID-19 [1, 11-18]. However, the main disadvantage of SIR based methods is that they require estimates of critical epidemiological parameters [19]. We developed a novel method to detect flattening and estimating the ending life-cycle of COVID-19 curves using only the time-series of New Cases Per Day (NCPD) which doesn’t require critical epidemiological parameters [20].

In this paper, an estimate using an optimistic scenario, based on the method [20], for the ending life-cycle and cumulative cases by the end of the major COVID-19 pandemic wave in India and some of its states is presented. An ideal scenario is also simulated, to provide hope and an opportunity to contain the COVID-19 outbreak at its earliest through appropriate policy interventions.

## 2. Methods

We used the method developed and reported in [20] to estimate the ending-life cycle and cumulative cases by the end of the major COVID-19 curve of India and some of its states. This method is explained briefly to maintain continuity. First, a time-series of New Cases Per Day (NCPD) is formed. The gross behaviour of the NCPD time-series is then fitted to a Gaussian function represented by the following equation:

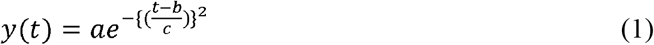

here, *t* is the time in days, and *y(t)* is the estimate of the gross behaviour of the NCPD time-series. We are considering only positive values of *t*. Coefficients *a, b* and *c* represent the amplitude of the peak, centroid (location), and width of the curve respectively. Coefficients of this equation are obtained as per the method [20] and then the estimation of the peak amplitude is made and the relative position of the peak is also obtained. A positive value of the relative peak position indicates, how far the peak is from the current day, while the negative value indicates that peak has already arrived. In this manner, using eqn.(1), ending life-cycle of the curve is also estimated. Here, ending life-cycle means the day when the curve will come to an end.

## 3. Data, simulation results, and discussion

We simulated, the NCPD time-series for India and some of the states contributing most of the NCPD of India on 22^nd^ May 2020, as tabulated in Table-1. We used data from [21], wherein the data is fetched from the website of the Ministry of Health & Family Welfare(MoHFW). All simulations consider the first day as the day when the relevant data was first reported on the website [21] i.e. 11^th^ March 2020. We divided our simulation results into the following scenarios as explained below:

**Table 1:** Coefficients *a, b*, and *c* for India and some of its states on 22^nd^ May 2020 using an optimistic scenario.

| Country/State | NCPD | $a$ | $b$ | $c$ |
| --- | --- | --- | --- | --- |
| India | 5871 | 8188.9 | 102.9 | 45.2 |
| Maharashtra | 2345 | 3623.6 | 102.7 | 41.1 |
| Tamil Nadu | 776 | 763.55 | 74.85 | 21.69 |
| Delhi | 571 | 662.93 | 95.82 | 43.09 |
| Gujarat | 368 | 495.27 | 75.18 | 32.73 |
| Uttar Pradesh | 340 | 251.30 | 102.97 | 55.06 |
| Bihar | 308 | 1111.11 | 102.8 | 22.4 |
| Madhya Pradesh | 246 | 274.89 | 103.31 | 55.82 |
| Rajasthan | 212 | 294.61 | 102.63 | 54.21 |

### Optimistic Scenario

This type of scenario is utilised when there is a consistent increase in NCPD with high values and thus leads to a situation, where projections of the COVID-19 curve parameters such as cumulative cases and ending life-cycle become too high. However, there is always a possibility that future data points may fall in values due to strict containment measures are taken in the recent past. To account for this fact, we allow dynamics of the COVID-19 curve to spread to the extent such that the estimates of these parameters do not take excessively high values in a relatively short span of time. In this situation, If we find that the relative position of the peak is having a positive value greater than 30 days, then we obtain modified coefficients *a, b*, and *c* such that the value of the relative position of the peak becomes equal to positive 30 days or nearest to it. Here, it is also important to note that very high values of such parameters should also be taken care of by the policy makers, if consistent with future data points.

### Ideal Scenario

This scenario, assumes that the NCPD will start decreasing within a few days and thus generate an estimate of the COVID-19 parameters in an ideal situation. This scenario allows policy makers to further tighten their containment measures. They can also get the idea regarding future estimates of these parameters and get motivated if data points fall according to the ideal scenario. Therefore, in the event when the relative position of the peak is having a positive value greater than 30 days, we obtain modified coefficients *a, b*, and *c* such that the value of the relative position of the peak becomes equal to positive one day or a value nearest to it. However, if we find that the relative position of the peak is having a negative value then we don’t obtain modified coefficients. After obtaining the modified or original coefficients, eqn. (1) is used to calculate cumulative cases and ending life-cycle using the COVID-19 curve. Here, we calculate ending dates when the estimated peak amplitude falls to value by 90% and 95 respectively. Table-1 shows the coefficients *a, b*, and *c* using optimistic or actual scenario and the NCPD values for India and some of its states.

### India

5871 new cases per day were reported in India on the 22^nd^ of May 2020. Figure 1(a) shows two estimates, one with higher projections of NCPD and another one with an optimistic scenario. With higher projections, the relative peak position is positive 44 days, while using the optimistic scenario, the relative position of the peak is positive 30 days. This can be seen in Fig.1 (a) where current position (in green color) is at the 73^rd^ day (22^nd^ May 2020) while the peak position (in red color) is at the 113^th^ and the 103^rd^ day for highly projected values of NCPD and optimistic scenario respectively. Estimate using higher projections of the NCPD indicates that the cumulative cases at the end of COVID-19 life-cycle may rise to 1056000 cases with ending dates for 90% and 95% of fall in the peak amplitude by the 21^st^ of September (1175 cases per day) and 30^th^ of September (587 cases per day) respectively. However, using the optimistic scenario, we obtain 655000 cumulative cases with ending dates for 90% and 95% of fall in the peak amplitude by the 29^th^ of August (818 cases per day) and 8^th^ of September (409 cases per day) respectively. Figure 1(b) shows the ideal scenario. To simulate this scenario, we assumed some low values of NCPD just after the 73^rd^ day. Such an estimate results in 267200 cumulative cases with ending dates for 90% and 95% of fall in the peak amplitude by the 15^th^ of July 2020 (474 cases per day) and 21^st^ of July 2020 (237 cases per day) respectively. The ideal scenario indicates that India may report 237 cases per day by the end of the third week of July 2020 provided appropriate and strict policy interventions are made.

**Figure. 1.**
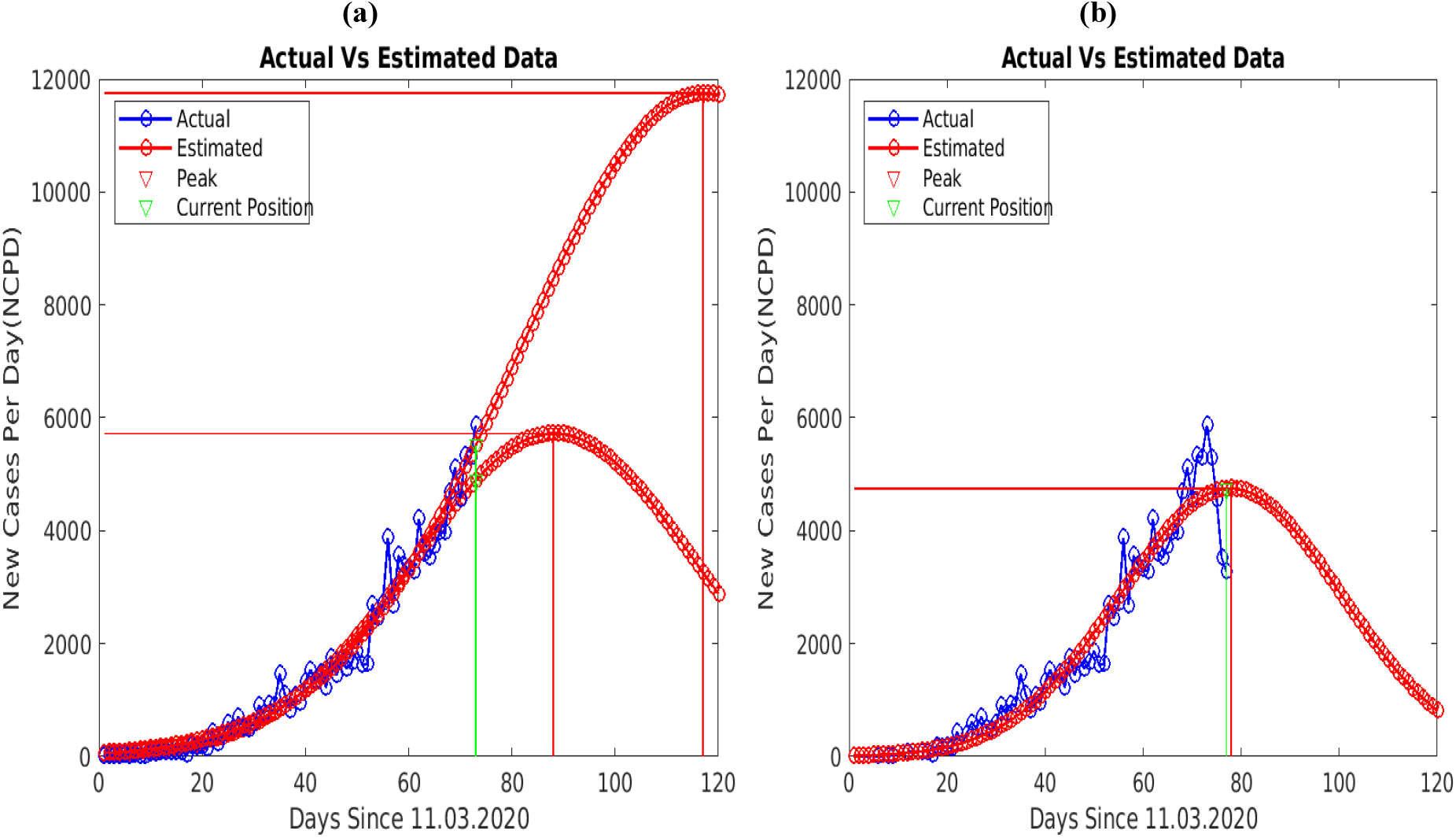
(a) Two estimates for India, one with higher projections of NCPD (upper-red) and another one with an optimistic scenario (lower-red), note that the position of the peak (vertical red line) with the higher projections is +44 days from the current position (vertical green line) while with optimistic scenario position of the peak is +30 days. (b) projections of NCPD with an ideal scenario

### Maharashtra

2345 new cases were reported in Maharashtra on the 22^nd^ of May 2020. Maharashtra contributed almost 40 % new cases per day for India on this day. The dynamics of COVID-19 curve for Maharashtra are shown in Fig. 2(a). As per the optimistic scenario, Maharashtra may report 263700 cumulative cases by the end of the COVID-19 life-cycle, while 362 and 181 new cases per day may be reported by the 22nd of August 2020 and 31^st^ of August 2020 respectively.

**Figure. 2.**
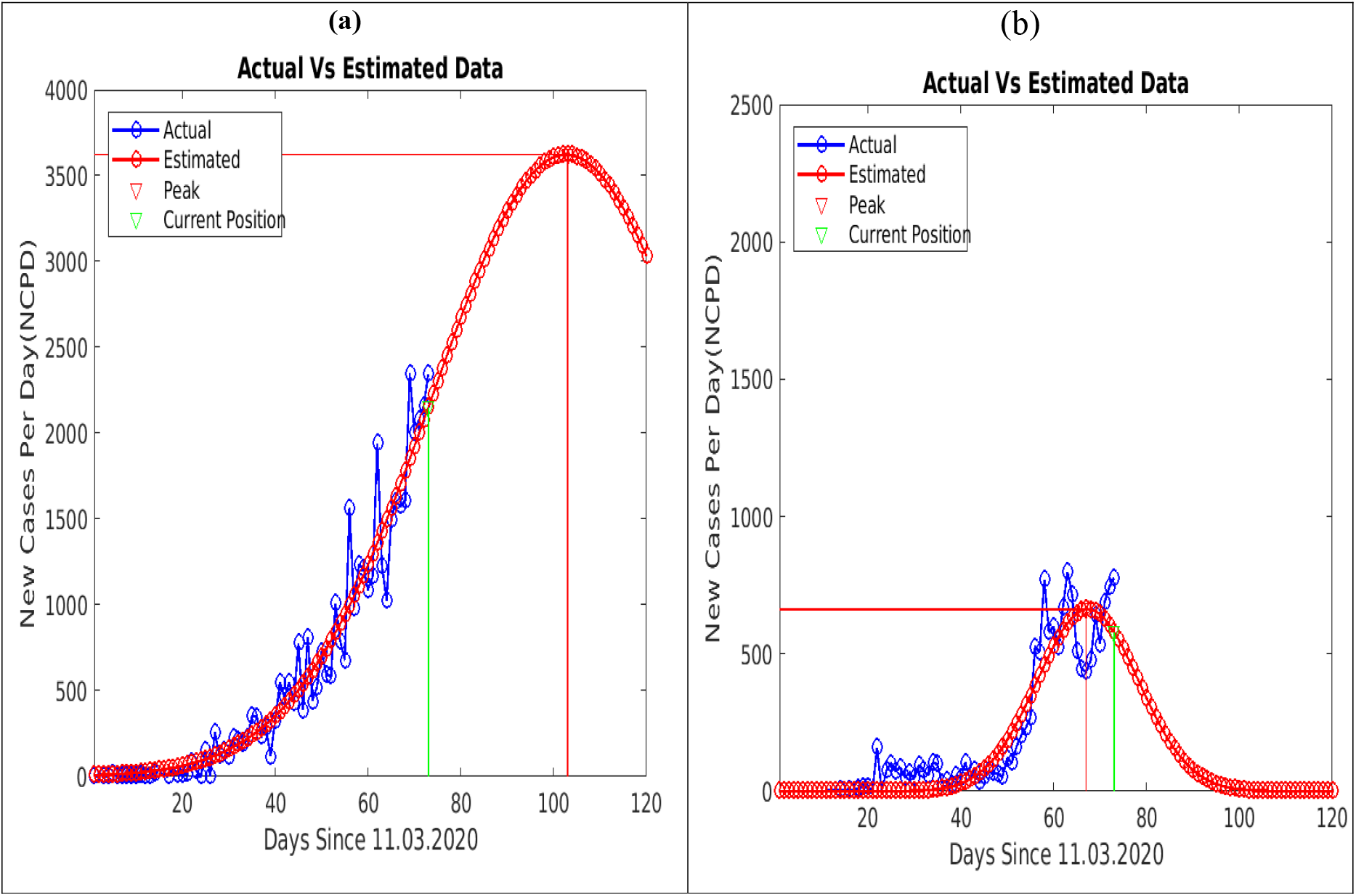
(a) Estimates using an optimistic scenario for Maharashtra, peak position (in red) is +30 days from the current position (in green) (b) Estimates using actual coefficients *a, b*, and *c* for Tamil Nadu, observe that the peak position is – 6 days from the current position.

### Tamil Nadu

776 new cases were reported in Tamil Nadu on the 22^nd^ of May 2020. The dynamics of the COVID-19 curve for Tamil Nadu are shown in Fig.2(b). Though Tamil Nadu reported the second-highest NCPD for India on the simulation day, yet the dynamics of the COVID-19 curve indicate flattening of the COVID-19 curve which is evident from the fact that the relative peak position is negative six days on the day of simulation. So, the Tamil Nadu Government has an opportunity to contain the outbreak by the 13^th^ of June 2020 with 33 new cases per day and 18140 cumulative cases by the end of COVID-19 curve. However, improper policy interventions may extend flattening of this curve with more cumulative cases.

### Delhi

571 new cases were reported in Delhi on the 22^nd^ of May 2020. The dynamics of COVID-19 curve for Delhi are shown in Fig. 3 (a). Simulation results suggest that, Delhi may report 50600 cumulative cases by the end of the COVID-19 life-cycle, while 66 and 33 new cases per day may be reported by the 19^th^ of August 2020 and 28^th^ of August 2020 respectively.

**Figure. 3.**
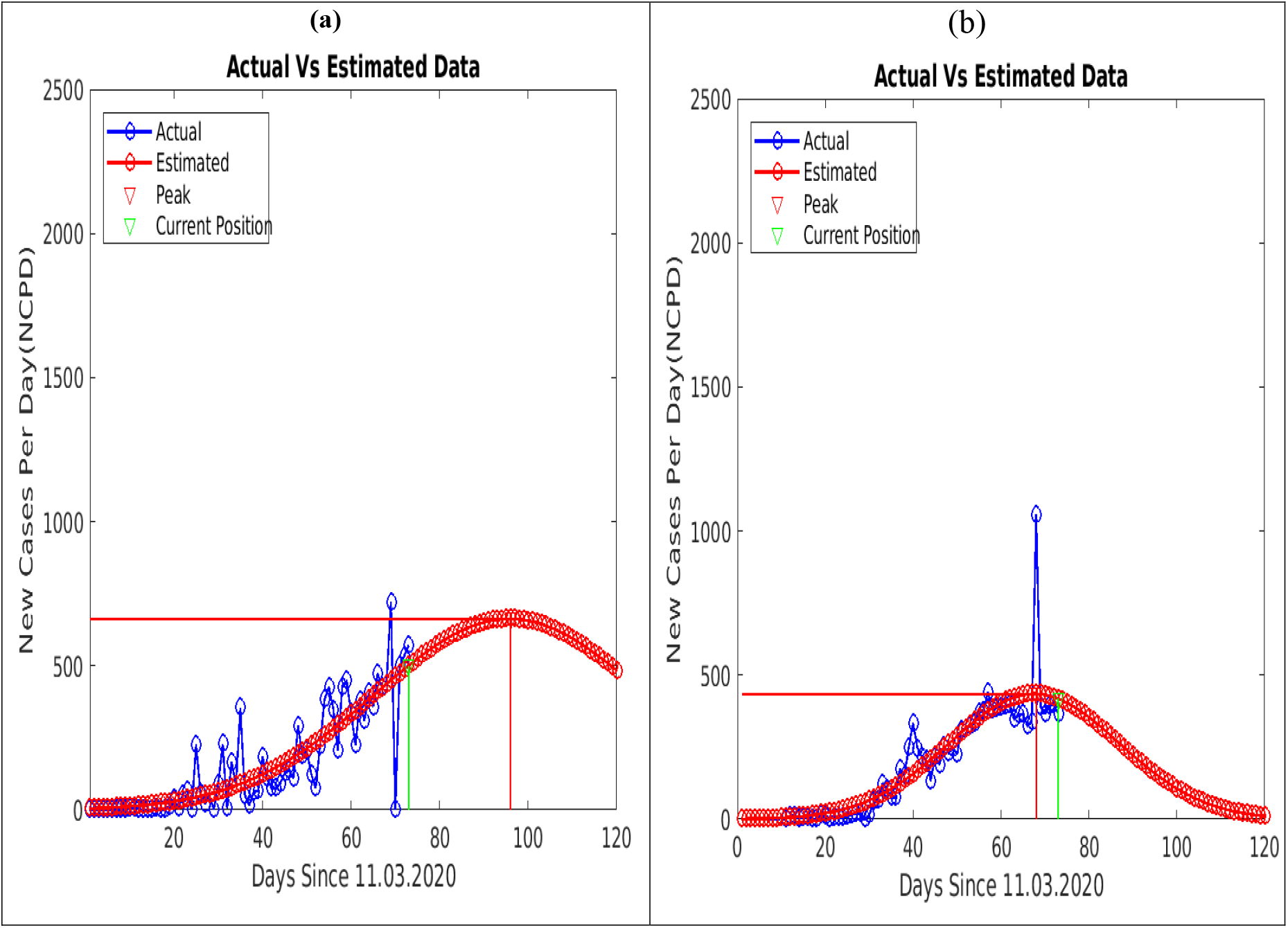
(a) Estimates using non-modified coefficients *a, b*, and *c* for Delhi, peak position (in red) is less than +30 days from the current position (in green) (b) Estimates using non-modified coefficients *a, b*, and *c* for Gujarat, note that the peak position is – 5 days from the current position, also observe that the COVID-19 curve for Gujarat started flattening.

### Gujarat

The dynamics of the COVID-19 curve for Gujarat is shown in Fig.3 (b). Gujarat reported forth highest NCPD for India on the simulation day. However, the dynamics of COVID-19 curve indicate flattening of the COVID-19 curve, which is evident from the fact that the relative peak position is negative five days on the day of simulation. So, there is an opportunity for the Gujarat Government to contain the outbreak by the 19^th^ of July 2020 with 33 new cases per day and 18140 cumulative cases by the end of the COVID-19 curve. However, improper policy interventions may also extend flattening of this curve with more cumulative cases.

### Uttar Pradesh

340 new cases were reported in Uttar Pradesh on the 22^nd^ of May 2020. The dynamics of the COVID-19 curve for Uttar Pradesh are shown in Fig.5(a). As per the optimistic scenario, Uttar Pradesh may report 24420 cumulative cases by the end of the COVID-19 life-cycle, while 25 and 12 new cases per day may be reported by the 13^th^ of September 2020 and 25^th^ of September 2020 respectively.

### Bihar

308 new cases were reported in Bihar on the 22^nd^ of May 2020. The dynamics of COVID-19 curve for Bihar are shown in Fig.4(b). As per the optimistic scenario, Bihar may report 44170 cumulative cases by the end of the COVID-19 life-cycle, while 111and 55 new cases per day may be reported by the 25^th^ of July 2020 and 29^th^ of July 2020 respectively. Our simulation results suggest that the Government of Bihar has performed well during initial period of pandemic and supressed the first small wave quite efficiently. Therefore, to simulate the second major wave, we didn’t consider the first small wave again. The big difference between ideal scenario and optimistic scenario is also an alarm to the Government of Bihar to lay down aggressive policy measures, to contain this outbreak

**Figure. 4.**
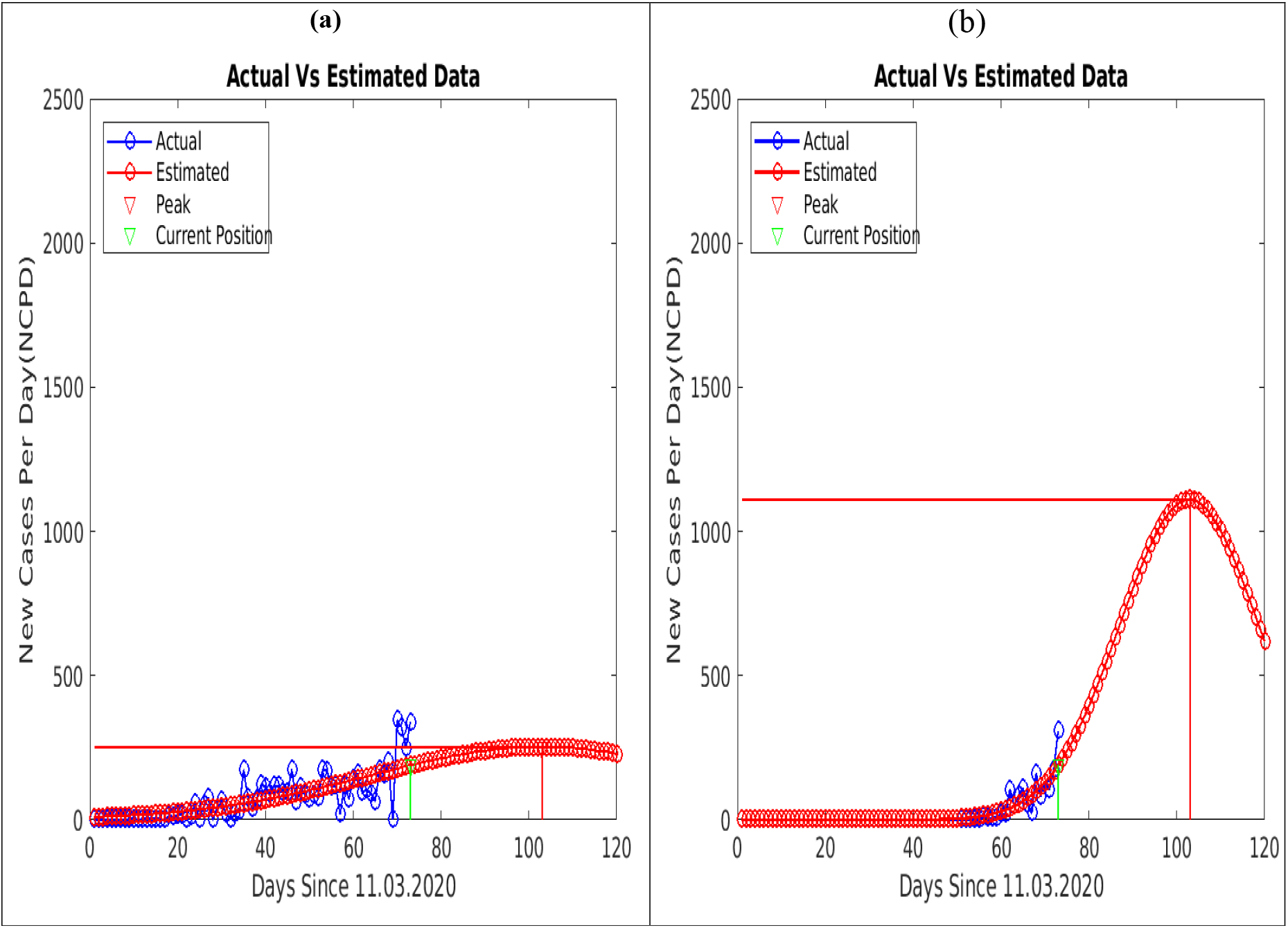
(a) Estimates using an optimistic scenario for Uttar Pradesh, note that position of the peak (in red) is +30 days from the current position (in green) (b) Estimates using an optimistic scenario for Bihar, note the higher peak amplitude.

### Madhya Pradesh

***246*** new cases were reported in Madhya Pradesh on the 22^nd^ of May 2020. The dynamics of the COVID-19 curve for Madhya Pradesh are shown in Fig.5(a). As per the optimistic scenario, Madhya Pradesh may report 27080 cumulative cases by the end of COVID-19 life-cycle, while 27 and 13 new cases per day may be reported by the 15^th^ of September 2020 and 26^th^ of September 2020 respectively.

**Figure. 5.**
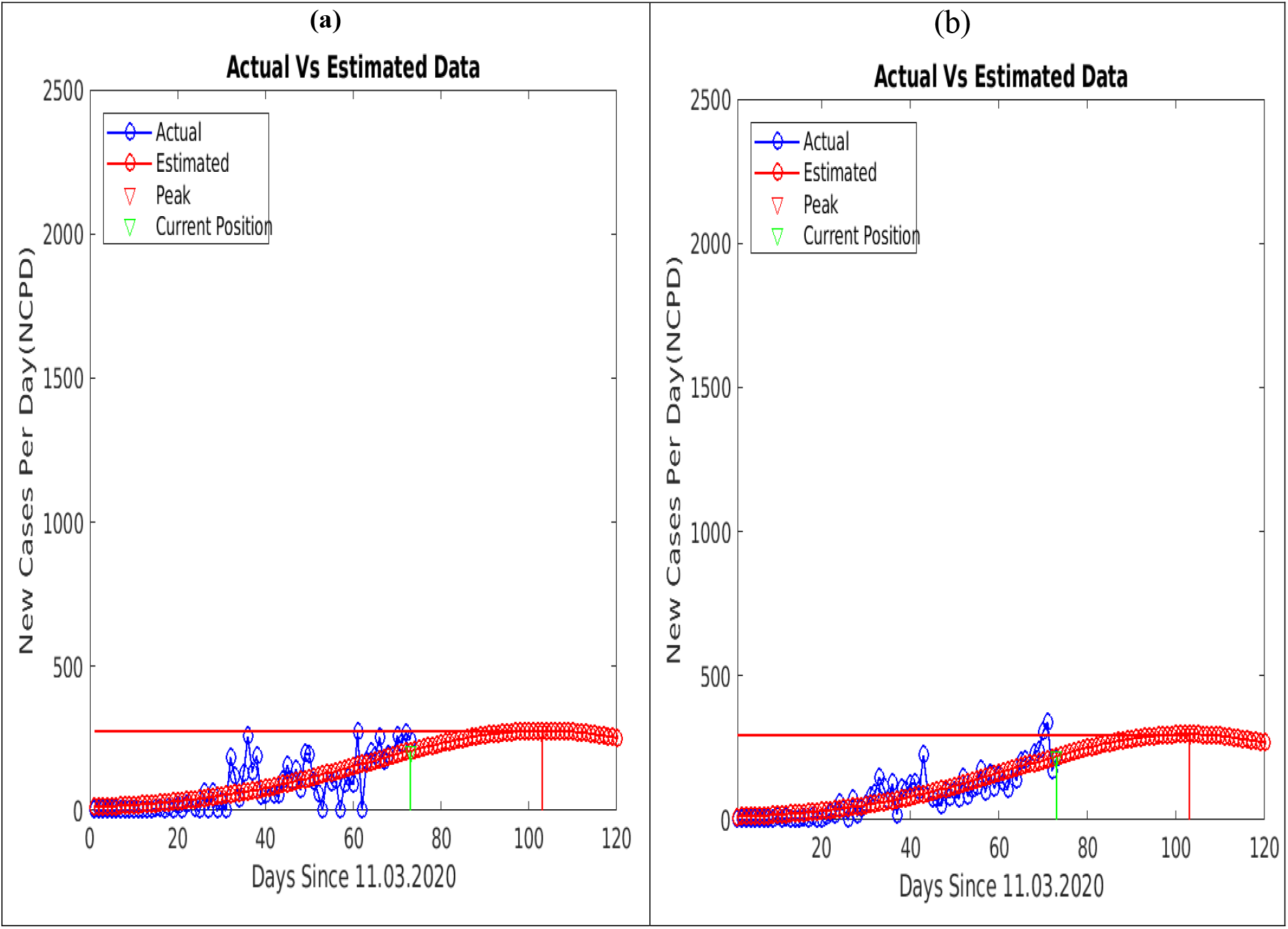
(a) Estimates using an optimistic scenario for Madhya Pradesh, note that the position of the peak (in red) is +30 days from the current position (in green) (b) Estimates using an optimistic scenario for Rajasthan. Note the prolonged flattening of pandemic waves in both the COVID-19 curves.

### Rajasthan

***212*** new cases were reported in Rajasthan on the 22^nd^ of May 2020. The dynamics of the COVID-19 curve for Rajasthan are shown in Fig.5(b). As per the optimistic scenario, Rajasthan may report 28200 cumulative cases by the end of COVID-19 life-cycle, while 29 and 14 new cases per day may be reported by the 15^th^ of September 2020 and 26^th^ of September 2020 respectively.

The Table 2, summarises the simulation results for COVID-19 parameters such as ending life-cycles and cumulative cases for India and some of its states in different scenarios.

**Table 2:** Estimates of cumulative cases and ending life-cycle for India and some of its states in different scenarios on the 22^nd^ of May 2020.

| Country/State | Scenario | Cumulative Cases by the end of the curve | *Fall of 95 % of the peak–amplitude by – 2020 and expected cases per day* |  | *Fall of 90 % of the peak–amplitude by – 2020 and expected cases per day* |  |
| --- | --- | --- | --- | --- | --- | --- |
| India | High | 1056000 | 30<sup>th</sup> September | 587 | 21<sup>st</sup> September | 1175 |
|  | Optimistic | 655000 | 8<sup>th</sup> September | 409 | 29<sup>th</sup> August | 818 |
|  | Ideal | 267200 | 21<sup>st</sup> July | 237 | 15<sup>th</sup> July | 474 |
| Maharashtra | Optimistic | 263700 | 31<sup>st</sup> August | 181 | 22<sup>nd</sup> August | 362 |
|  | Ideal | 92190 | 13<sup>th</sup> July | 94 | 7<sup>th</sup> July | 188 |
| Tamil Nadu | Actual/Ideal | 18140 | 13<sup>th</sup> June | 33 | 9<sup>th</sup> June | 66 |
| Delhi | Actual | 50600 | 28<sup>th</sup> August | 33 | 19<sup>th</sup> August | 66 |
|  | Ideal | 23970 | 19<sup>th</sup> July | 21 | 12<sup>th</sup> July | 42 |
| Gujarat | Actual/Ideal | 21130 | 3<sup>rd</sup> July | 22 | 28<sup>th</sup> June | 44 |
| Uttar Pradesh | Optimistic | 24420 | 25<sup>th</sup> September | 12 | 13<sup>th</sup> September | 25 |
|  | Ideal | 13110 | 11<sup>th</sup> August | 8 | 02<sup>nd</sup> August | 17 |
| Bihar | Optimistic | 44170 | 29<sup>th</sup> July | 55 | 25<sup>th</sup> July | 111 |
|  | Ideal | 3060 | 12<sup>th</sup> June | 8 | 10<sup>th</sup> June | 16 |
| Madhya Pradesh | Optimistic | 27080 | 26<sup>th</sup> Sept. | 13 | 15<sup>th</sup> September | 27 |
|  | Ideal | 13930 | 09<sup>th</sup> August | 9 | 31<sup>st</sup> July | 19 |
| Rajasthan | Optimistic | 28200 | 23<sup>rd</sup> Sept. | 14 | 11<sup>th</sup> September | 29 |
|  | Ideal | 14220 | 07<sup>th</sup> August | 10 | 29<sup>th</sup> July | 20 |

## 4. Conclusion

Overestimating the COVID-19 pandemic wave parameters such as its ending life-cycle and cumulative cases may put policy makers in a dilemma. In this paper, an optimistic scenario is simulated, wherein the dynamics of the COVID-19 curve is allowed to spread to such an extent that the projections of the COVID-19 parameters do not take excessively high values. This study, suggests that by the end of major COVID-19 wave in India, there may be 655000 cumulative cases which may not be considered too high in a population of 1.38 billion people. Maharashtra is one of the major contributing states of the Indian COVID-19 curve. Pandemic waves in Tamil Nadu and Gujarat are in the flattening phase. The state government of Bihar requires special attention to contain COVID-19 outbreak while Delhi, Utter Pradesh, Madhya Pradesh, and Rajasthan may end-up with prolonged flattening.

## Data Availability

we obtained the data from the labnol website. The data is fetched from the MoHFW website.

https://www.labnol.org/code/covid-19-india-tracker-200325

